# Prevalence and causes of hospitalization-related sarcopenia: A scoping review

**DOI:** 10.1101/2025.03.22.25324464

**Authors:** Yoshinori Yamamoto, Masato Ogawa, Takayuki Okamoto, Marika Tsuboi, Ryo Momosaki

## Abstract

**Objectives:** Sarcopenia is characterized by the progressive loss of muscle mass and strength and poses significant health risks, particularly for older adults. Recent studies have focused on hospitalization-related sarcopenia—a rapid decline in muscle mass and strength during hospitalization—that has emerged as a critical prognostic factor. This screening review aimed to examine the prevalence of hospitalization-related sarcopenia and risk factors.

**Methods:** This scoping review was conducted following the PRISMA-ScR guidelines. A literature search was performed in MEDLINE and the Cochrane Central Register of Controlled Trials in December 2022. Studies on patients who developed sarcopenia because of hospitalization were included. Meta-analyses, reviews, and nonprimary studies were excluded, with a focus on selecting cohort studies. The risk of bias was assessed using the risk-of-bias assessment tool for nonrandomized studies.

**Results:** Of the 775 articles screened, 9 were included. Studies from eight countries showed hospitalization-related sarcopenia prevalence of 14%–75%. Major risk factors included low skeletal muscle mass index, low body mass index, and length of hospitalization. Acute illness and surgery significantly accelerated sarcopenia, with rapid muscle mass loss observed within days to weeks of bed rest. Prolonged bed rest, systemic inflammation, and malnutrition due to hypermetabolism were identified as major contributors to muscle mass loss during hospitalization.

**Conclusions:** Hospitalization-related sarcopenia occurs in a short period after surgery or acute illness. This review highlights the need for awareness of hospitalization-related sarcopenia and the importance of developing strategies to reduce its effect.

## 1. Introduction

Sarcopenia is a condition characterized by gradually decreasing muscle mass and strength, posing important health risks, particularly for older adults. Sarcopenia has a significant effect on physical function and quality of life. Muscle mass typically declines by 0.1%–0.5% per year with aging. ^1,2)^ However, muscle mass can be reduced by factors other than age. In recent years, the concept of “hospitalization-related sarcopenia,” a rapid loss of muscle mass and strength during hospitalization, has gained attention.^3)^

Hospitalization is associated with several factors that accelerate the progression of sarcopenia. Extended bed rest, inflammation, and nutritional deficiencies are key contributors to muscle mass loss. ^4,5)^ For instance, older adults experience substantial losses in muscle mass and strength after just 10 days of bed rest. ^6,7)^ Moreover, severe illness can trigger acute inflammation, accelerating muscle mass loss by increasing the production of inflammatory cytokines and disrupting muscle protein metabolism. ^3,8)^ Despite recognizing these risks, the specific mechanisms that accelerate hospitalization-related sarcopenia are not well understood. Studies have reported the general prevalence of sarcopenia^9–11)^; however, very few studies have focused on hospitalization-related sarcopenia.^2)^ The existing literature^12,13)^ lacks detailed longitudinal studies on hospitalization-related sarcopenia and the effect of acute factors.

This review investigated how hospitalization accelerates the progression of hospitalization-related sarcopenia. Understanding the causes of this type of sarcopenia is crucial for preventing muscle loss during hospitalization. This scoping review aimed to determine the prevalence and causes of hospitalization-related sarcopenia, synthesize current knowledge, and contribute to the development of clinical research.

## 2. Material and methods

This scoping review was carried out in accordance with the guidelines of the Preferred Reporting Items for Systematic Reviews and Meta-Analyses Extension for Scoping Reviews. ^14)^

### 2.1 Search Strategy

MEDLINE and the Cochrane Central Register of Controlled Trials were searched for literature in December 2022. The search strategies were initially developed through team discussions and were further refined with the assistance of a librarian. These strategies combined keywords related to both sarcopenia and hospitalization modalities. Detailed information on the search strategies employed is available in Table 1.

**Table 1.**
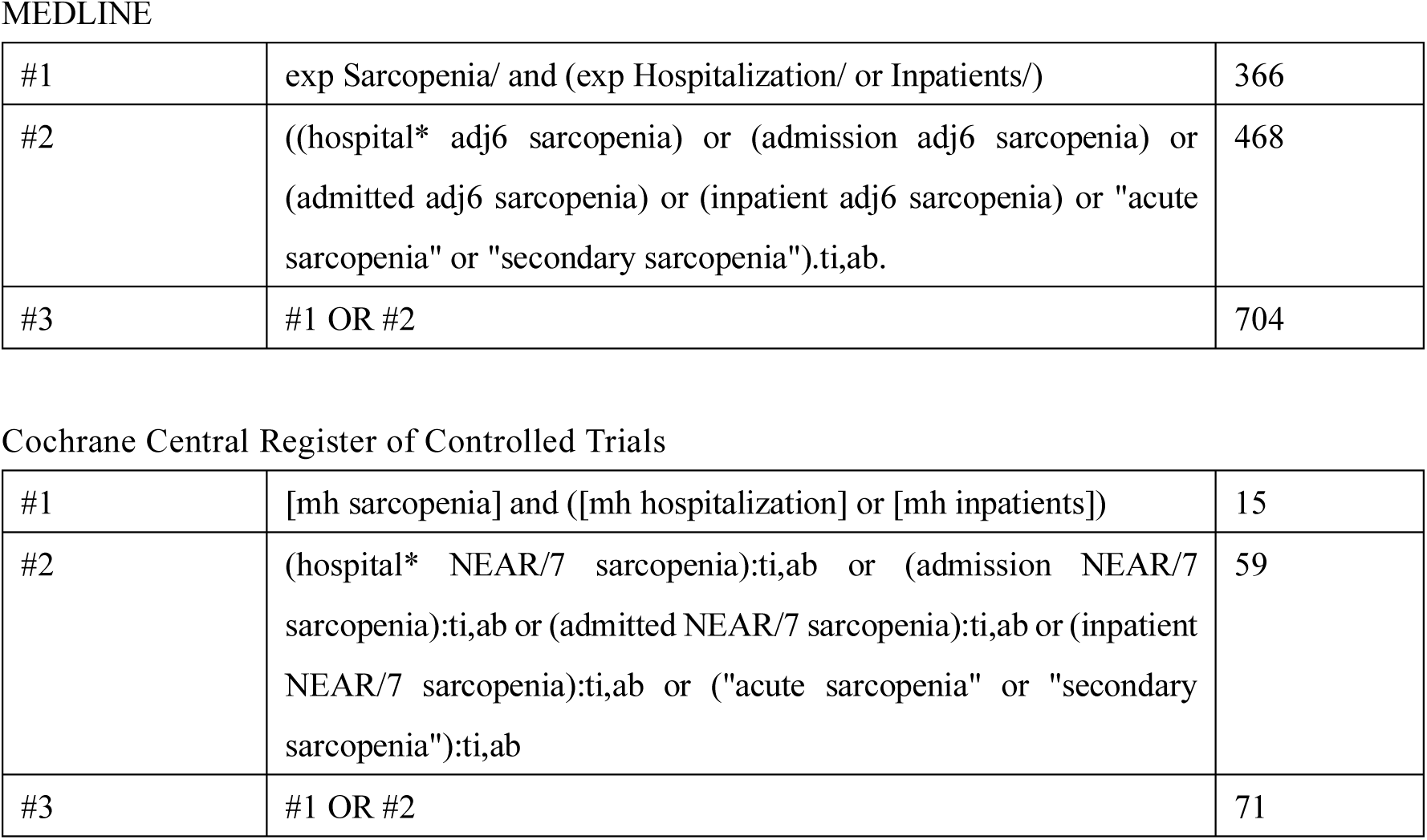
The final search string for MEDLINE and the Cochrane Central Register of Controlled Trials.

|  |  |  |
| --- | --- | --- |
| #1 | exp Sarcopenia/ and (exp Hospitalization/ or Inpatients/) | 366 |
| #2 | ((hospital* adj6 sarcopenia) or (admission adj6 sarcopenia) or (admitted adj6 sarcopenia) or (inpatient adj6 sarcopenia) or "acute sarcopenia" or "secondary sarcopenia").ti,ab. | 468 |
| #3 | #1 OR #2 | 704 |
| Cochrane Central Register of Controlled Trials |  |  |
| #1 | [mh sarcopenia] and ([mh hospitalization] or [mh inpatients]) | 15 |
| #2 | (hospital* NEAR/7 sarcopenia):ti,ab or (admission NEAR/7 sarcopenia):ti,ab or (admitted NEAR/7 sarcopenia):ti,ab or (inpatient NEAR/7 sarcopenia):ti,ab or ("acute sarcopenia" or "secondary sarcopenia"):ti,ab | 59 |
| #3 | #1 OR #2 | 71 |

### 2.2 Inclusion Criteria and Exclusion Criteria

Each team member reviewed articles published in English from database inception to December 2022 for eligibility. The inclusion criteria were as follows: patients who developed sarcopenia during hospitalization, specifically those who were evaluated for sarcopenia after admission and subsequently received a definitive diagnosis. Sarcopenia was diagnosed based on the criteria established by several leading organizations: European Working Group on Sarcopenia in Older People (EWGSOP)^2)^, Asian Working Group for Sarcopenia (AWGS)^15)^, International Working Group on Sarcopenia^16)^, and Foundation for the National Institutes of Health.^17)^ In addition, cases of presarcopenia—defined as the loss of muscle mass without a concomitant decline in muscle strength or physical function—and assessments made using the SARC-F screening tool^18)^ were also enrolled. This study followed a cohort design.

The exclusion criteria were as follows: patients who had already been diagnosed with sarcopenia before or at the time of admission, as well as meta-analyses, reviews, systematic reviews, and letters to the editor.

### 2.3 Data Extraction and Synthesis

To determine eligibility, the four researchers were divided into two groups, and each group independently assessed the eligibility of the articles. Each of the two reviewers within the groups independently evaluated eligibility based on study characteristics (authors, study design, country, and duration of study) and description of the study population (age, sex, and disease status). Studies were selected by (1) title screening, (2) abstract review, and (3) full-text assessment. Any disagreements were resolved through discussion with the other research team members. The potential for bias was evaluated utilizing the risk-of-bias assessment tool specifically designed for non-randomized^19)^

## 3. Results

### 3.1 Selection Process

The database search yielded 775 candidate articles for the scoping review. After removing 74 duplicates, 701 titles and abstracts were screened based on the inclusion and exclusion criteria. This step identified 33 studies that met the inclusion criteria, and their full texts were reviewed. Of these articles, 24 were excluded based on the exclusion criteria, leaving 9 articles eligible for the scoping review (Figure 1).

**Figure 1.**
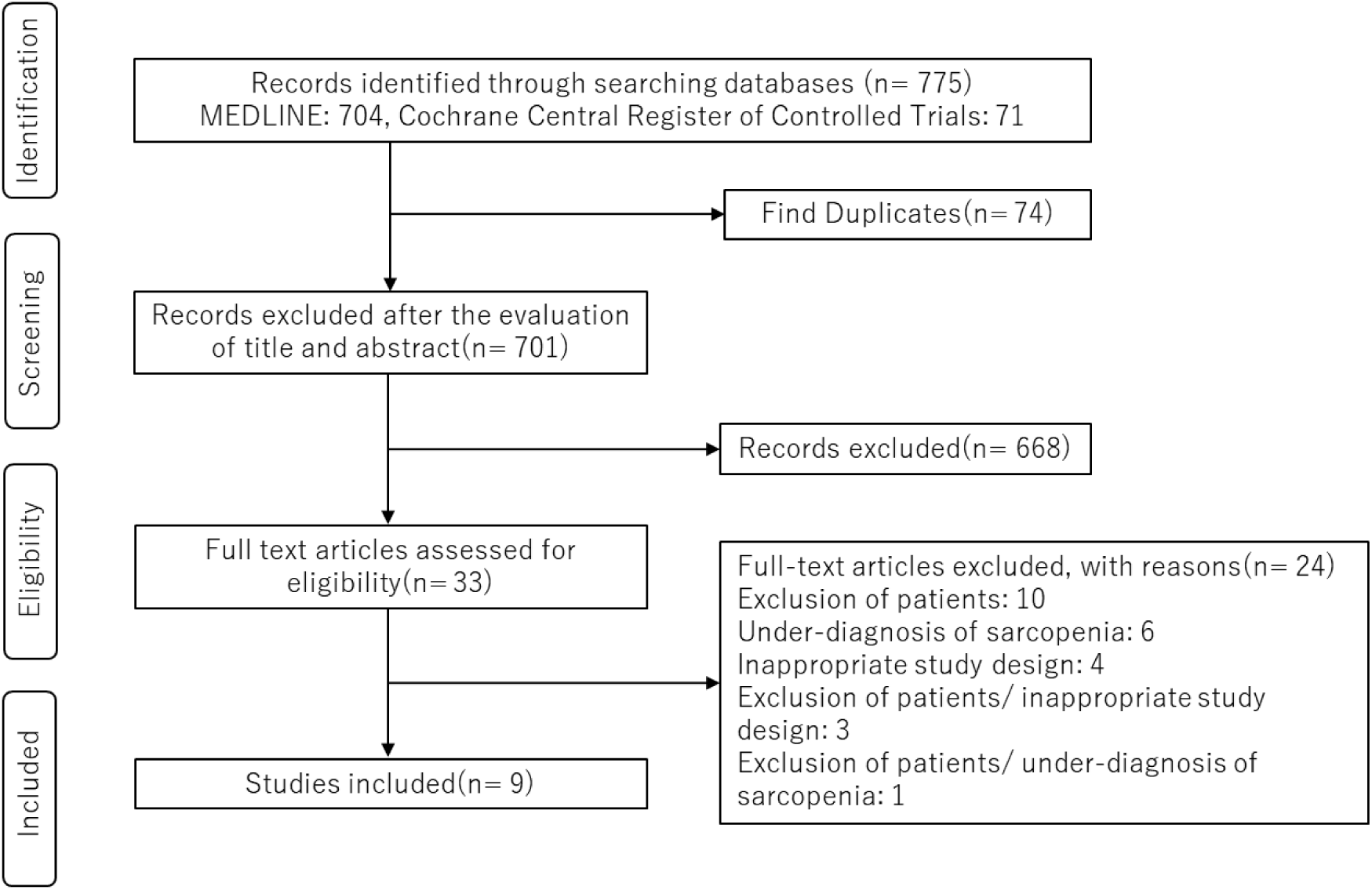
PRISMA flow diagram.

### 3.2 Overview of Included Studies

Table 2 presents an overview of the study characteristics. The studies originated from eight countries: 1 from Canada^20)^, 3 from the UK^21–23)^, 1 from Italy^24)^, 1 from the USA^25)^, 1 from Spain^26)^, 1 from Thailand^27)^, and 1 from France. ^28)^ The studies included 6 cases involving surgery^20,22,23,25–27)^, 1 case undergoing cancer treatment^21)^, 1 case in geriatric medicine^26)^, and 1 case with COVID-19.^28)^

**Table 2.**
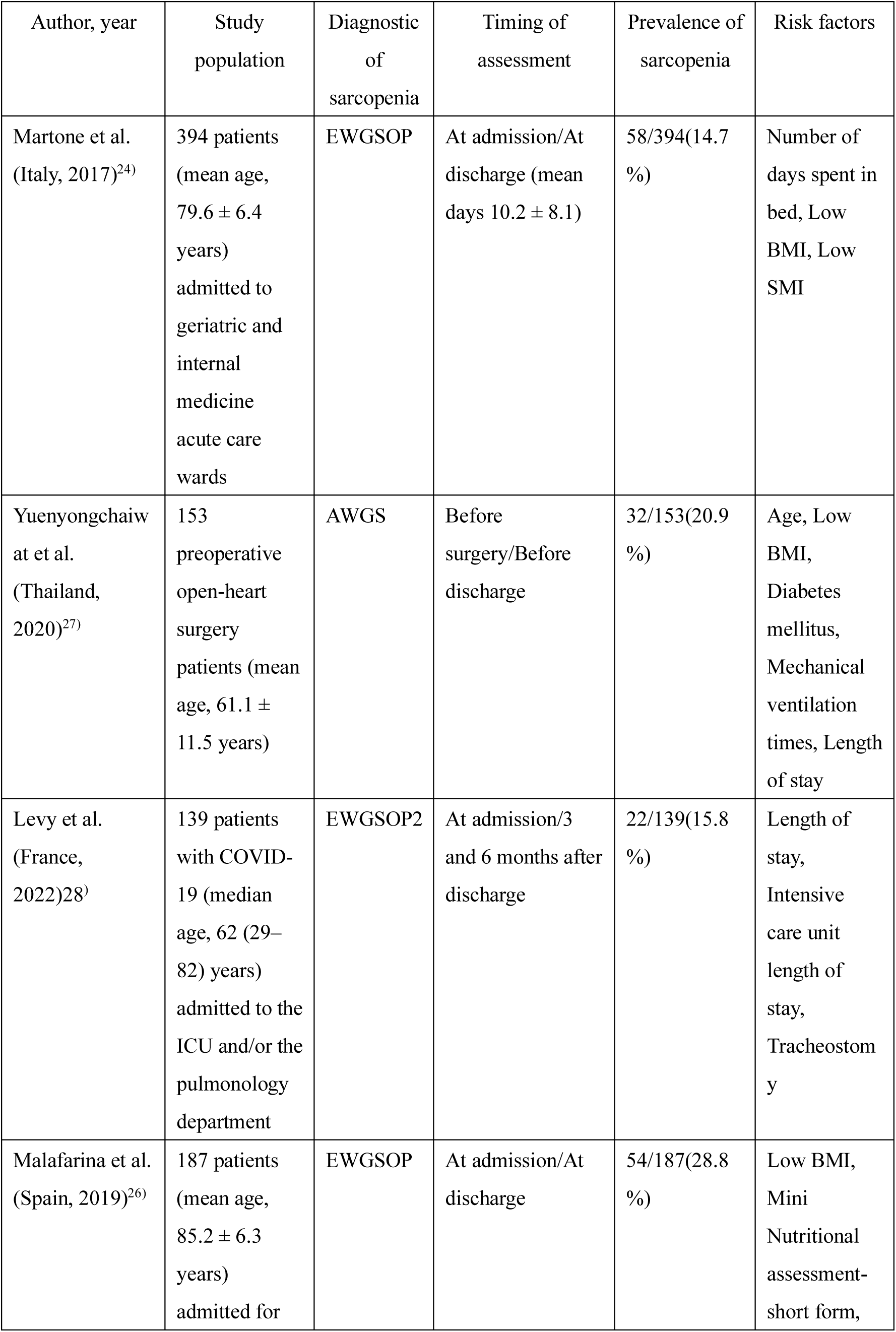
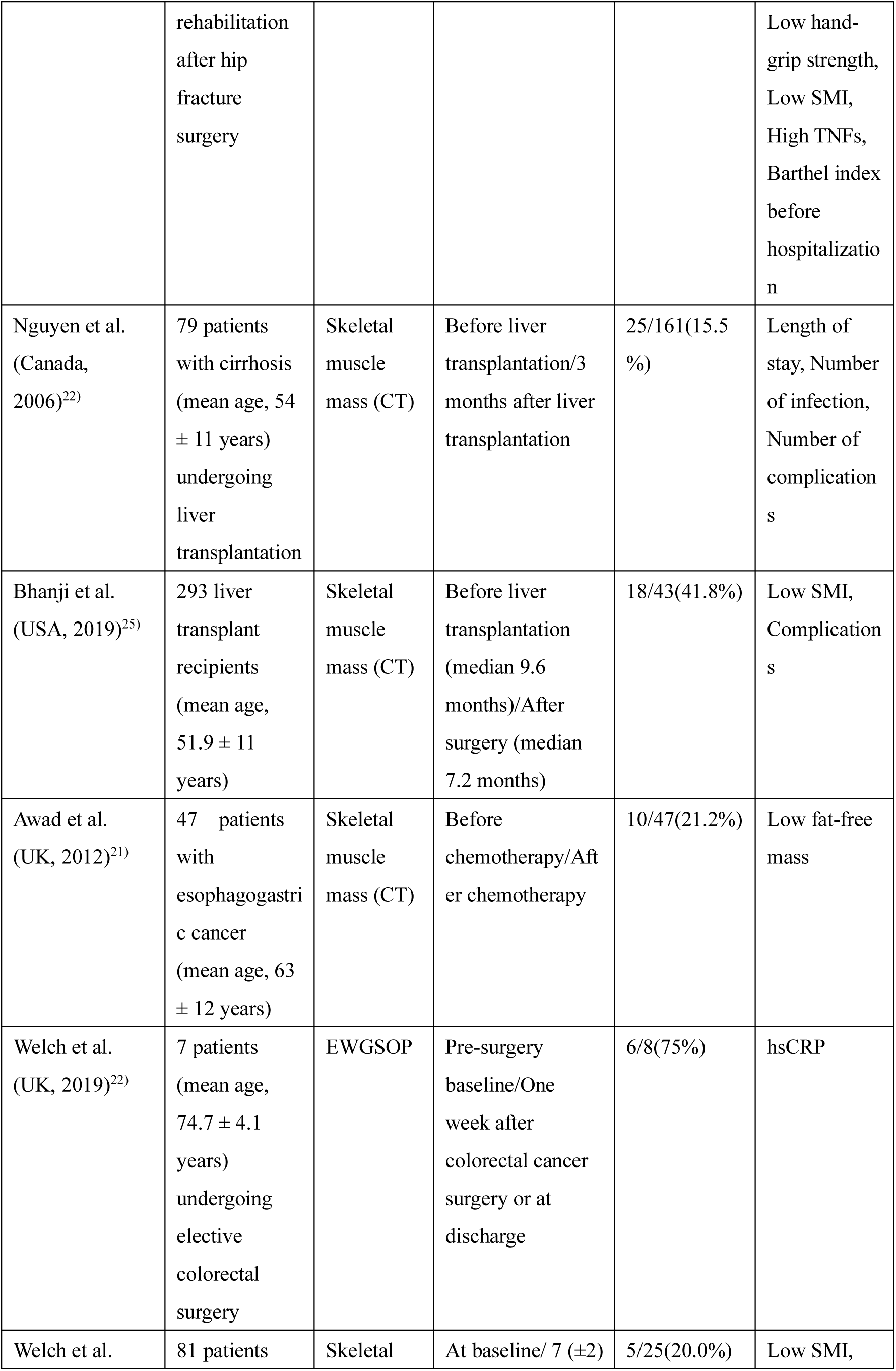
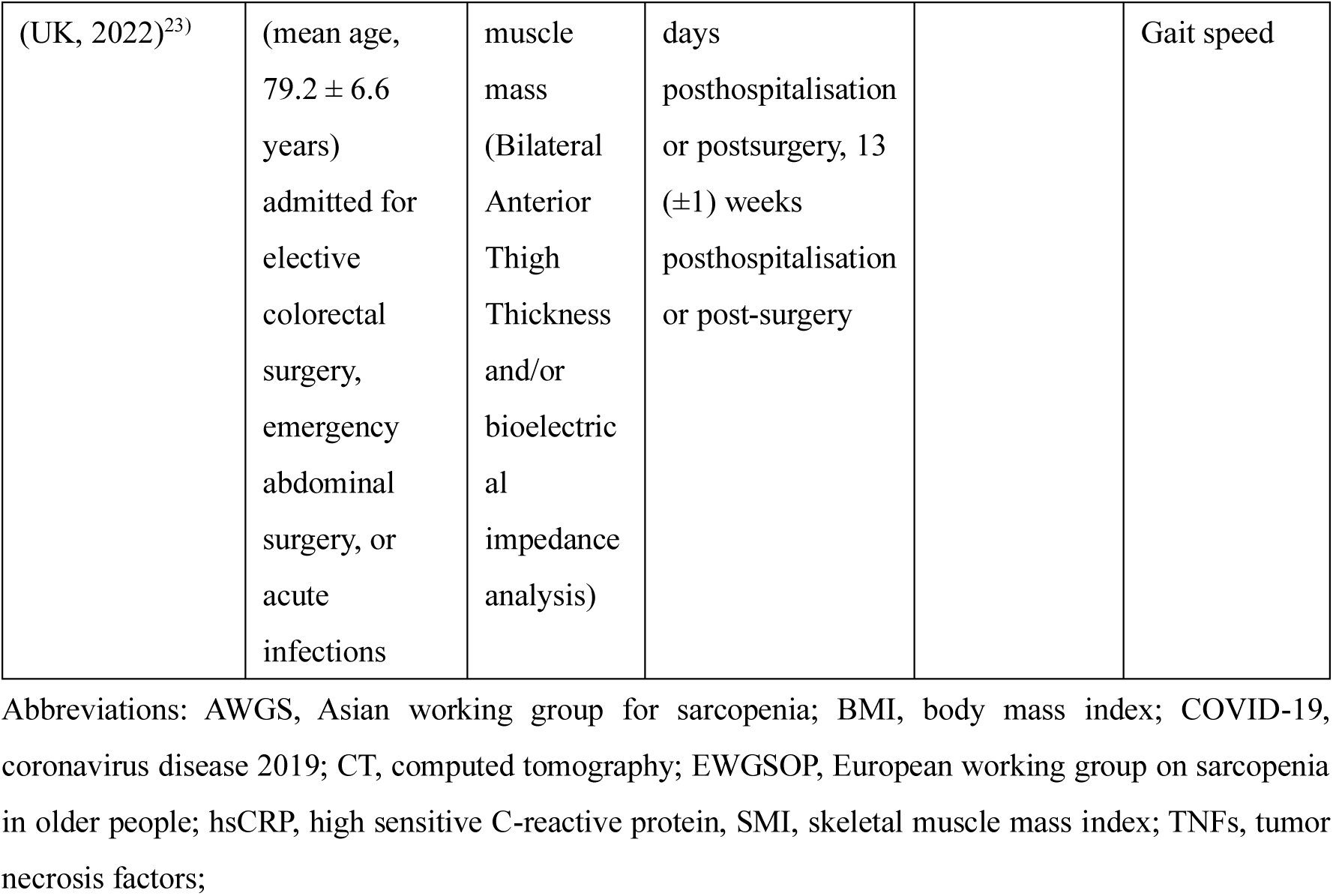
Characteristics of the included studies.

### 3.3 Diagnostic criteria and incidence of sarcopenia and risk factors

Diagnostic criteria for sarcopenia were as follows: three studies used the EWGSOP criteria^22,24,26)^, one used the EWGSOP2 criteria^28)^, one used the AWGS criteria^27)^, and four used the criteria based on skeletal muscle mass alone.^20,21,23,25)^

The prevalence of sarcopenia is presented in Figure 2 Sarcopenia developed in 14%–75% of patients during hospitalization or within months after discharge.^20–28)^ Some studies also reported proportions >40%^22,25)^; however, these studies involved smaller sample sizes.

**Figure 2.**
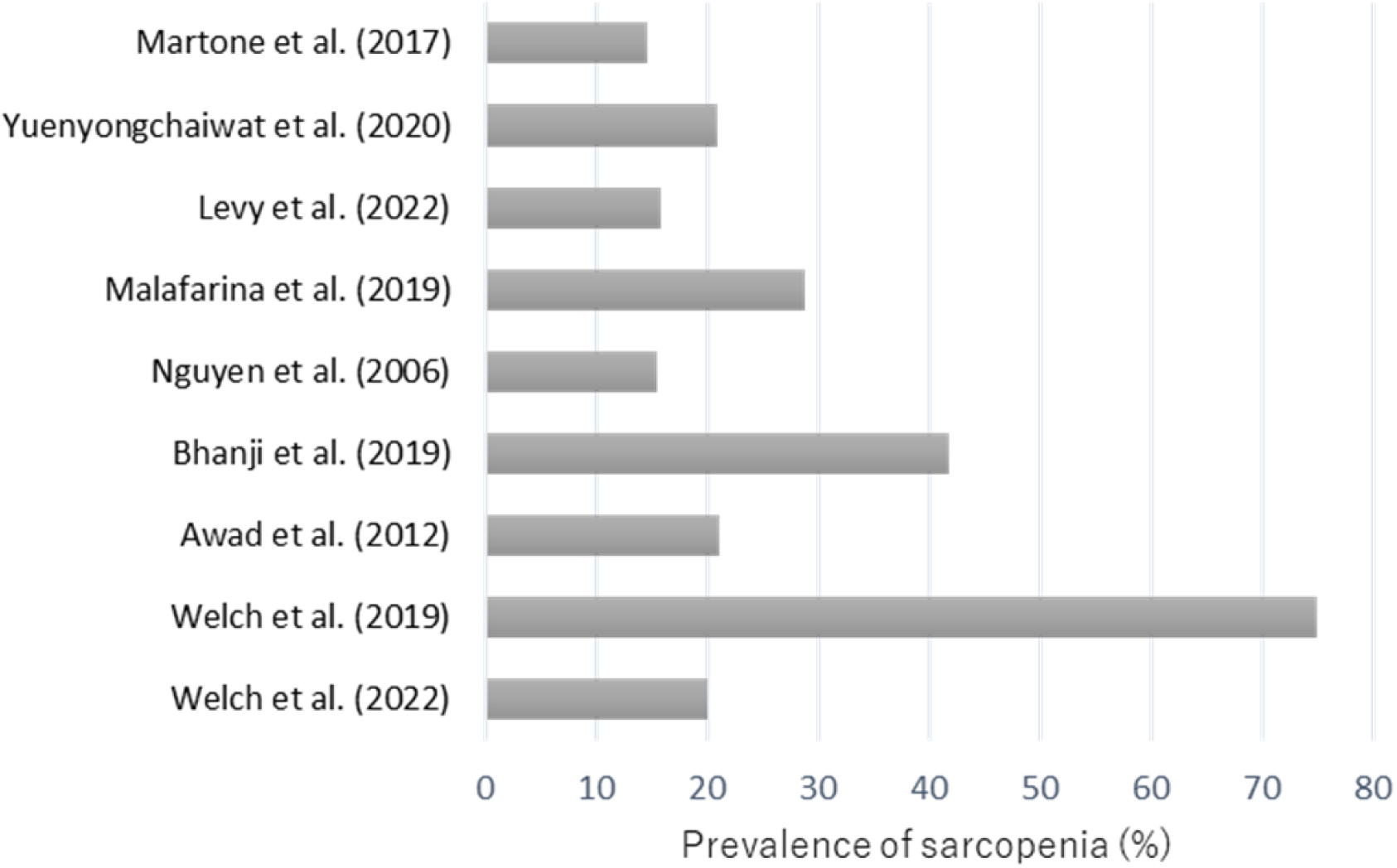
Prevalence of sarcopenia in different studies.

Risk factors are provided in Table 3. Risk factors for sarcopenia development included low skeletal muscle mass index (SMI) in three studies^24–26)^, low body mass index (BMI) in three^24,26,27)^ and length of stay in three.^20,27,28)^

**Table 3.**
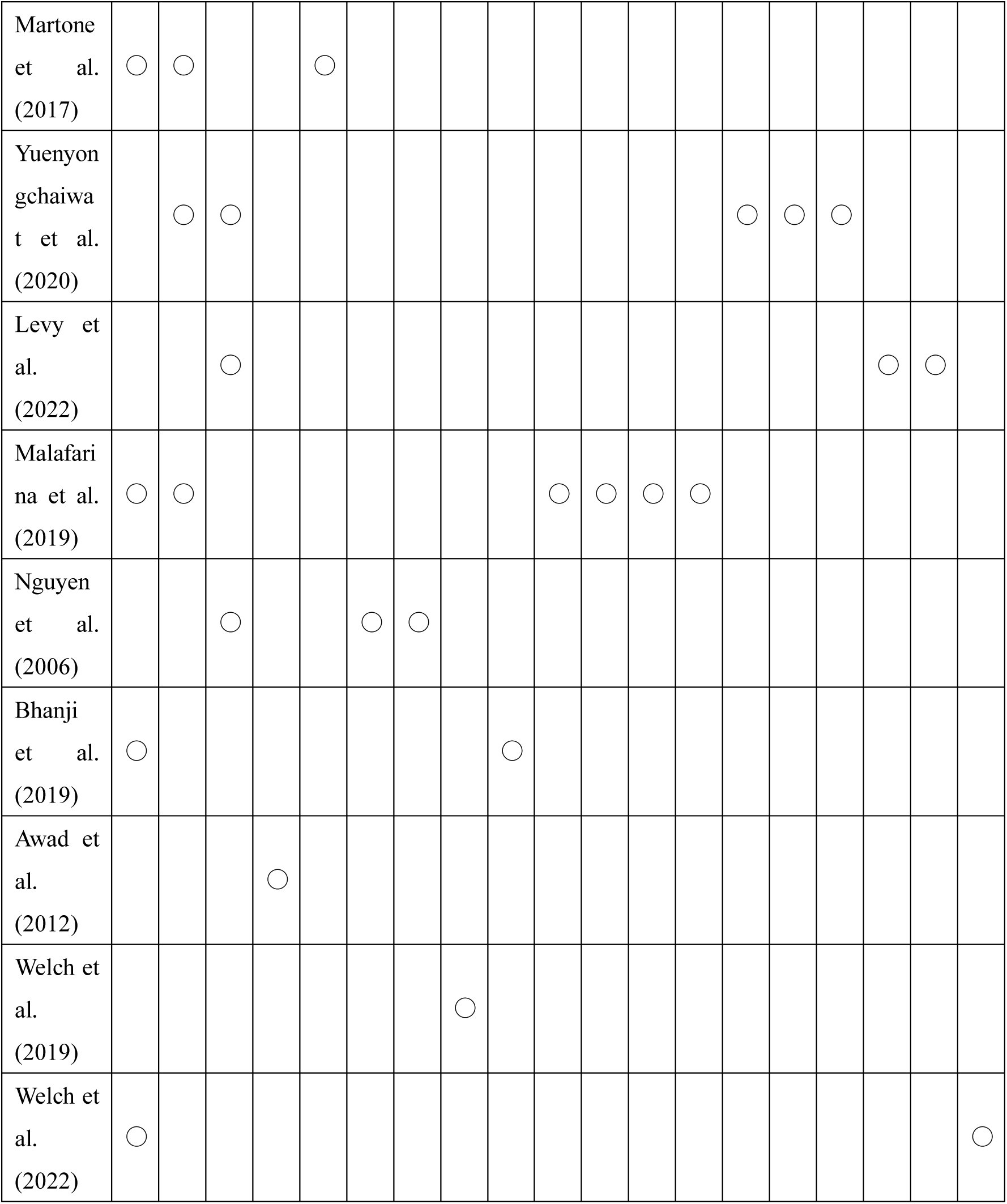
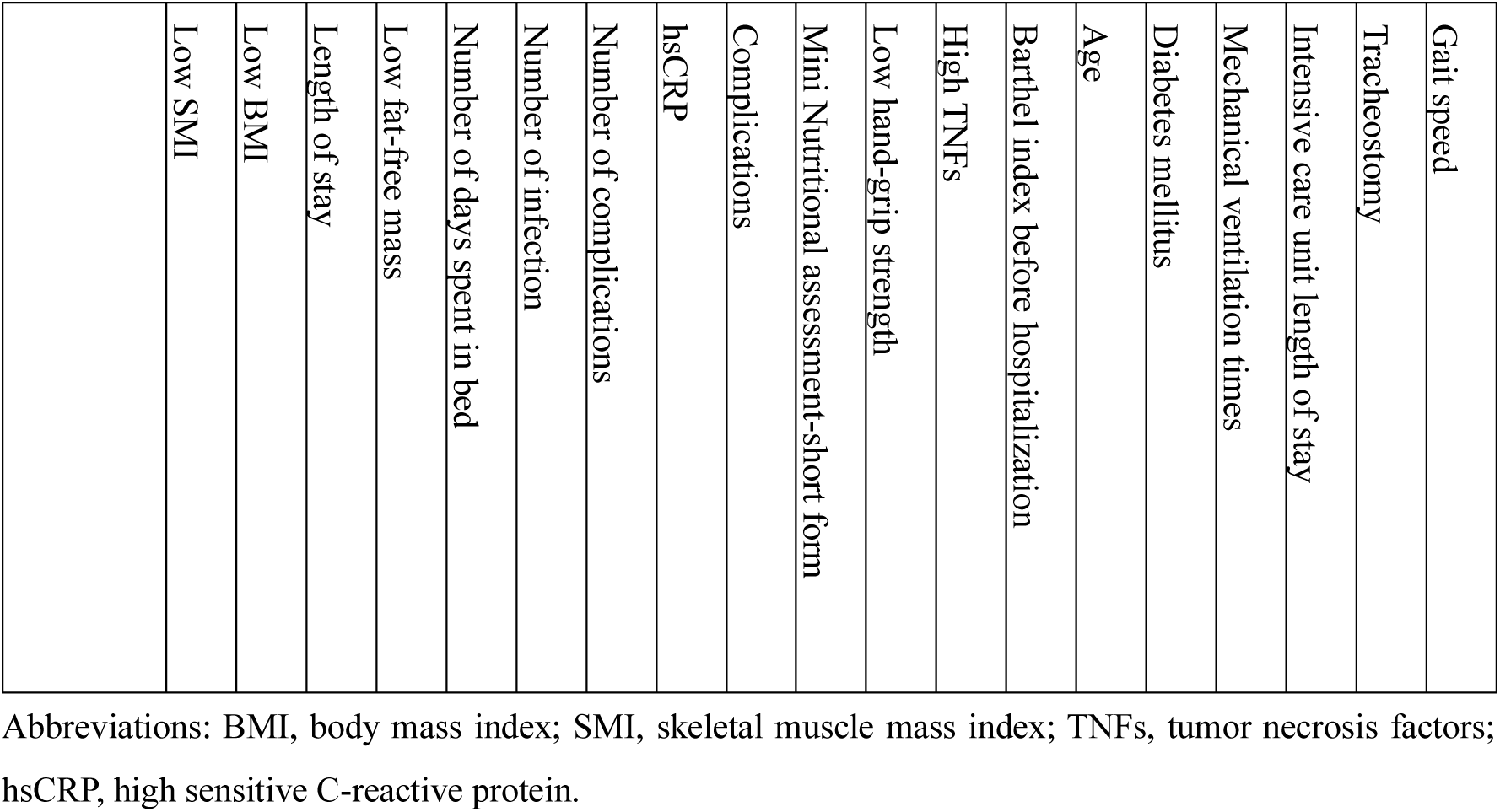
Risk factors of hospitalization-related sarcopenia in each study.

### 3.4 Risk-of-bias assessment

The results of the risk-of-bias assessment are presented in Figure 3. Most studies (8 out of 9, 89%) were rated as low risk in the participant selection domain. Similarly, 6 out of 9 (67%) studies were rated as low risk in both the confounding variables and exposure measurement domains. The highest frequency of high-risk assessments was observed in the outcome assessment blinding and incomplete outcome data domains, with 6 out of 9 (67%) studies rated as high risk. In addition, the selective outcome reporting domain was rated as high risk in 5 out of 9 (56%) studies. Unclear risk was identified in the confounding variables, exposure measurement, and outcome assessment blinding domains, with 1–3 (11%–33%) studies rated as unclear risk.

**Figure 3.**
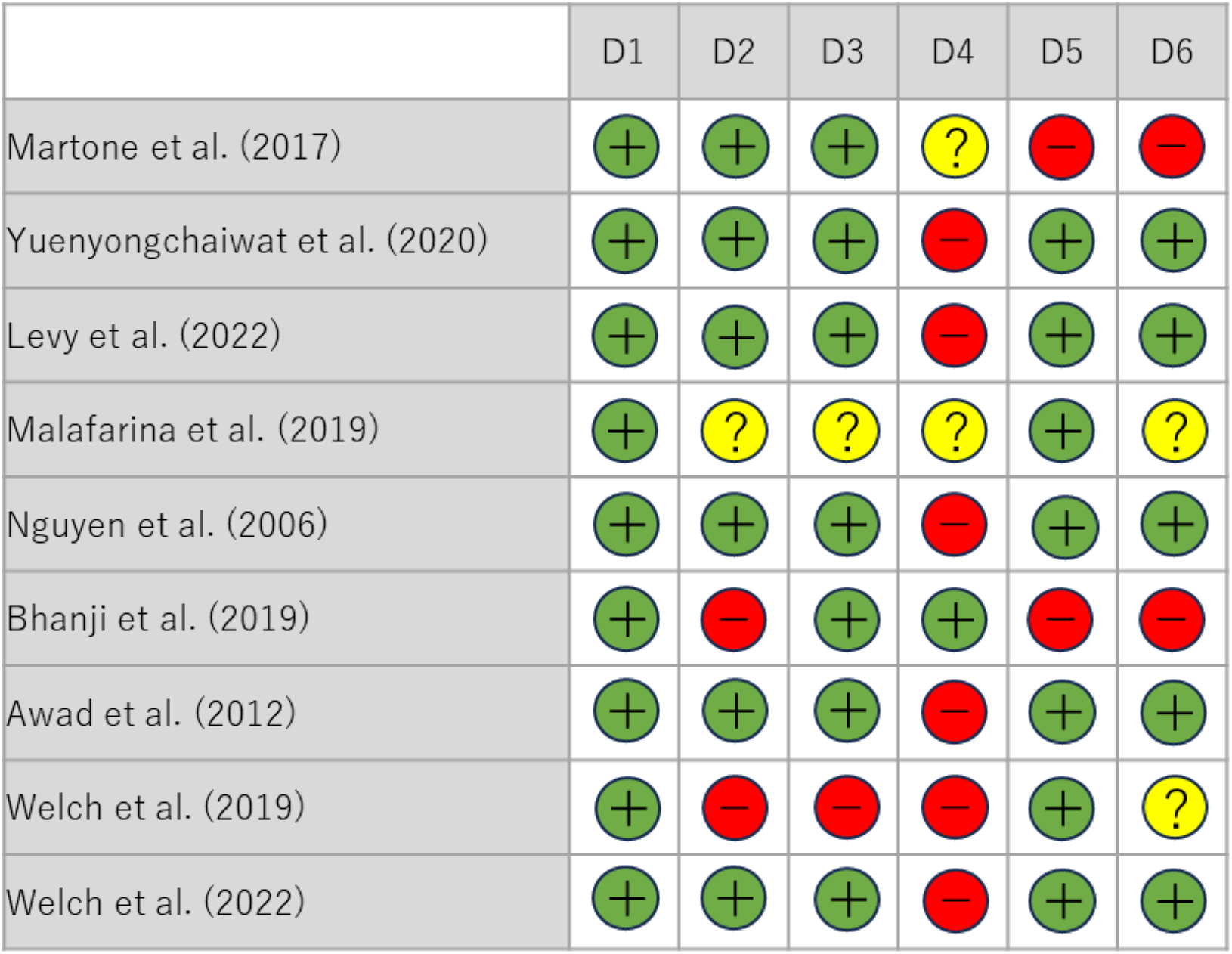
Risk-of-bias assessment. Domains: D1, selection of participants; D2, confounding variables; D3, measurement of exposure; D4, blinding of outcome assessments; D5, incomplete outcome data; D6, selective outcome reporting. + low risk, − high risk,unclear.

## 4. Discussion

This review highlights the potential for hospitalization-related sarcopenia to develop in a short period. In recent years, research and interest in hospitalization-related sarcopenia have increased. This review provides useful information to help healthcare professionals understand the risks of hospitalization-related sarcopenia and take steps to prevent its onset.

The results of this review indicate that hospitalization-related sarcopenia occurs frequently. Previous studies have examined the prevalence of sarcopenia in community-dwelling older adults, hospitalized patients, and patients with different conditions^9–11)^; however, data on hospitalization-related sarcopenia have been limited. This review revealed that hospitalization-related sarcopenia developed in 14%–75% of patients, which is higher than the global prevalence of sarcopenia of 10%–27% among individuals aged ≥65 years.^29)^ Certain disease groups have been reported to have a particularly high prevalence of sarcopenia, for example, 43.6% of patients with gastrointestinal cancer^30)^, 40.7% of patients who liver transplantation^31)^, and 48.0% in patients with COVID-19.^32)^ In these disease groups, systemic inflammation, physical inactivity, and undernutrition increase the risk of sarcopenia. Thus, this review presents that patients with such diseases are at increased risk of developing hospitalization-related sarcopenia.

Moreover, this review shows that acute illness and surgery increase the risk of hospitalization-related sarcopenia. In particular, bed rest rapidly reduces muscle mass in a short period. For example, muscle strength and lean body mass are significantly reduced after 5 days of bed rest in older adults^7)^, and muscle fiber cross-sectional area is significantly reduced after 14 days of bed rest in middle-aged adults.^33,34)^ Even healthy older adults lose at least 10% of muscle mass and up to 13% of knee extension muscle strength after 7–10 days of bed rest.^7,35,36)^ This review reveals that sarcopenia developed within 7–10 days of evaluation.^21–23)^ These results reflect that acute activity restriction is associated with a high risk of sarcopenia in the short term. Furthermore, acute illness and surgery trigger a systemic inflammatory response that accelerates muscle proteolysis. The increased production of cytokines associated with the inflammatory response leads to decreased muscle mass and function.^37)^ In addition, increased amino acid consumption and decreased albumin levels due to hypermetabolism lead to malnutrition.^38–40)^ A previous review reported observation of similar effects during renal dysfunction, chemotherapy, after liver transplantation, and during rehabilitation after a hip fractur.^25–27)^ After surgery, activity limitations because of pain and stress cause a rapid decrease in muscle mass and strength.^21)^ Activity restrictions during hospitalization and prolonged bed rest due to COVID-19 also contribute to the development of a systemic inflammatory response and undernutrition.^28)^ These findings demonstrate that risk factors associated with acute illness and surgery may cause acute sarcopenia. Particularly in older adults, these factors result in the rapid loss of muscle mass and strength, significantly increasing the risk of sarcopenia.

The strength of this review is attributed to its comprehensive synthesis of research on hospitalization-related sarcopenia. This review is based on a search strategy using MEDLINE and the Cochrane Central Register of Controlled Trials. Despite variations in the diagnostic criteria of sarcopenia, this review is an important resource of data that can improve our understanding of the prevalence and risk factors of hospitalization-related sarcopenia.

However, this review has several limitations. First, the small sample size of the studies reviewed, coupled with differences in their quality, could impact the reliability of the results. ^11)^ Secondly, the diagnostic criteria for sarcopenia differ among studies, leading to inconsistent findings and emphasizing the importance of standardization. ^9)^ Third, the high risk of blinding of outcome assessments and incomplete outcome data in this study raises concerns about the effect of bias. Fourth, the high risk of selective outcome reporting also contributes to the unreliability of the results. Fifth, unclear risks in some areas may limit the generalizability of the study results. Sixth, whether sarcopenia develops during hospitalization or after discharge remains unclear. Finally, this review included sarcopenia assessments after hospital discharge but was unable to control for factors such as post-discharge living conditions.^20,25,28)^

## 5. Conclusion

This review examined the prevalence and risk factors of hospitalization-related sarcopenia. Owing to the lack of adequate data in the existing literature, this review clarified these issues. The results indicate that the prevalence of hospitalization-related sarcopenia ranges from 14% to 75%, and it occurs within a short period after surgery or acute illness. Future longitudinal and prospective studies should be pursued to identify factors that contribute to the development of hospitalization-related sarcopenia. In addition, a comprehensive study of disease, duration, and treatment due to hospitalization is warranted to clarify the association between sarcopenia and hospitalization. Further studies are needed to examine long-term outcomes and the effects of early interventions such as exercise therapy, nutritional supplementation, and use of novel therapeutic targets.

## Data Availability

MEDLINE and the Cochrane Central Register of Controlled Trials

## Acknowledgement

We are grateful to Nara Medical University Library, the librarian from the Takaki Oseto, for her technical and invaluable assistance in the development and validation of the search strategy. We would like to extend our sincere gratitude to the members of the 2024 clinical practice guidelines committee for rehabilitation nutrition for their invaluable support.

